# The acceptance of covid-19 vaccines in Sub-Saharan Africa: Evidence from 6 national phone surveys

**DOI:** 10.1101/2021.06.28.21259320

**Authors:** Shelton Kanyanda, Yannick Markhof, Philip Wollburg, Alberto Zezza

## Abstract

**Introduction:** Recent debates surrounding the lagging covid-19 vaccination campaigns in low-income countries center around vaccine supply and financing. Yet, relatively little is known about attitudes towards covid-19 vaccines in these countries and in Africa in particular. In this paper, we provide cross-country comparable estimates of the willingness to accept a covid-19 vaccine in six Sub-Saharan African countries.

**Methods:** We use data from six national high-frequency phone surveys from countries representing 38% of the Sub-Saharan African population (Burkina Faso, Ethiopia, Malawi, Mali, Nigeria, and Uganda). Samples are drawn from large, nationally representative sampling frames providing a rich set of demographic and socio-economic characteristics by which we disaggregate our analysis. Using a set of re-calibrated survey weights, our analysis adjusts for the selection biases common in remote surveys.

**Results:** Acceptance rates in the six Sub-Saharan African countries studied are generally high, with at least four in five people willing to be vaccinated in all but one country. Vaccine acceptance ranges from nearly universal in Ethiopia (97.9%, 97.2% to 98.6%) to below what would likely be required for herd immunity in Mali (64.5%, 61.3% to 67.8%). We find little evidence for systematic differences in vaccine hesitancy by sex or age but some clusters of hesitancy in urban areas, among the better educated, and in richer households. Safety concerns about the vaccine in general and its side effects emerge as the primary reservations toward a covid-19 vaccine across countries.

**Conclusions:** Our findings suggest that limited supply, not inadequate demand, likely presents the key bottleneck to reaching high covid-19 vaccine coverage in Sub-Saharan Africa. To turn intent into effective demand, targeted communication campaigns bolstering confidence in the safety of approved vaccines and reducing concerns about side effects will be crucial to safeguard the swift progression of vaccine rollout in one of the world’s poorest regions.

What is already known?

- Estimates of vaccine acceptance in high- and middle-income countries have found rates to cluster around 70% with considerable cross-country variation.
- As vaccine rollout is severely lagging, low-income countries and particularly Sub-Saharan Africa (SSA) with almost two thirds of the global poor population remain exposed to covid-19 and its impacts on lives and livelihoods.
- Much of the current debate on vaccination campaigns in SSA focuses on supply-chain and financial factors, yet there is a dearth of robust, comparable evidence on covid-19 vaccine hesitancy in these countries.

What are the key findings?

- Covid-19 vaccine acceptance is high in six Sub-Saharan African countries with an estimated four in five people or more in all but one study country willing to take an approved, free vaccine.
- Clusters of hesitancy vary by country but generally comprise urban areas, richer households, and those with more education.

What do the new findings imply?

- Limited supply, rather than inadequate demand, likely presents the key bottleneck to achieving high covid-19 vaccine coverage in Sub-Saharan Africa.
- To reach mass coverage, information campaigns should bolster confidence in vaccine safety and alleviate concerns about side effects.

## Introduction

As vaccination campaigns in high-income countries are accelerating, large swathes of the global population living in low- and middle-income countries remain severely exposed to the novel coronavirus disease (covid-19).[1] Sub-Saharan Africa is home to 433 million people living below the international absolute poverty line (about two thirds of the global poor population).[2] For these populations, non-pharmaceutical interventions to curb the spread of the disease impinged on already precarious livelihoods.[4] Furthermore, the region is characterized by high prevalence of comorbidities, and public health systems that are ill equipped to stave off the burden of mass infection.[1,3,4] Reaching large-scale vaccination coverage in Sub-Saharan Africa (SSA) is thus pivotal in the global effort to halt the spread of the disease and limit its toll on lives and livelihoods.[5–7]

Recent calls to action by international stakeholders and a growing body of scholarly research focus on supply-chain and financial factors to safeguard sufficient vaccine availability in SSA. The COVAX initiative aims to provide doses sufficient to vaccinate up to 20% of the population in the region, far below the target population required for a coverage of 60%, or 321.5 million individuals.[8,9] Still, this would leave an estimated financing gap of over $10 billion in SSA.[10] A June 2021 resolution from the G7 pledging one billion covid-19-vaccine doses for low-income countries offers hope for those supply gaps being filled. While these considerations concern the availability of vaccines, another key factor for the success of vaccination campaigns in SSA is the willingness to be vaccinated within the population. Yet, there is a dearth of large-scale evidence on vaccine acceptance in low income countries in general and SSA in particular.

Based on a cross-country comparable sample with national scope from six Sub-Saharan African countries, we fill this knowledge gap by providing estimates of vaccine acceptance for a population representative of around 416 million people, 38% of the population of Sub-Saharan Africa.[11] Drawing on the high frequency phone surveys (HFPS) based on pre-pandemic sampling frames from nationally representative, face-to-face household surveys supported by the World Bank’s Living Standard Measurement Study – Integrated Survey on Agriculture (LSMS-ISA), we are able to link covid-19 vaccine acceptance rates to a rich set of demographic and socio-economic characteristics. Along with recalibrated sampling weights, this allows our study to provide robust insights into the likelihood of the current efforts to ensure sufficient supply of vaccination doses to also meet adequate demand in SSA, identify clusters of hesitancy, and contribute to a swift rollout of vaccination campaigns where they will be needed most.

## Methods

### Data and survey instrument

We use data from the national longitudinal high-frequency phone surveys on covid-19 (HFPS) conducted in Burkina Faso, Ethiopia, Malawi, Mali, Nigeria, and Uganda. Survey implementation was led by the respective national statistical agencies, the Burkina Faso National Institute of Statistics and Demography; Laterite Ethiopia in collaboration with the Central Statistical Agency of Ethiopia; Malawi National Statistical Office; Mali National Institute of Statistics; Nigeria Bureau of Statistics; and Uganda Bureau of Statistics; and supported by the World Bank Living Standards Measurement Study and the Poverty and Equity Global Practice. The HFPS on covid-19 have been implemented monthly since May 2020, aiming to gauge the impact of the covid-19 pandemic on individual and household attitudes, socioeconomic and health outcomes.[12]

The HFPS rounds in which vaccine hesitancy was covered in a cross-country comparable fashion were conducted in September 2020 (Ethiopia, round 6), October-November 2020 (Malawi, round 5; Mali, round 5; and Nigeria, round 6) and December 2020 (Burkina Faso, round 5; and Uganda, round 4). The primary survey questions of interest, posed to each phone survey respondent, was: ‘*If an approved vaccine to prevent coronavirus was available right now at no cost, would you agree to be vaccinated?’* with response options ‘*yes*’, ‘*no*’, and ‘*not sure’*;. If respondents answered ‘*no*’ or *‘not sure’*, this was followed up by a question about possible reasons for refusing to be vaccinated: ‘*What are the reasons you would not agree to be vaccinated?’*, with response options:

1. *‘I don’t think it will be safe’*
2. *‘I am worried about the side effects’*
3. *‘It is against my religion’*
4. *‘I am not at risk of contracting covid-19’*
5. *‘I don’t think it will work’*
6. *‘I am against vaccine in general’*
7. *Other*

We are interested in the association between willingness to be vaccinated and a standard set of demographic and socioeconomic variables, such as gender, age, income, and education. These are available for respondents either from the HFPS directly or from pre-covid-19 face-to-face (FtF) surveys, which served as the sampling frames for the HFPS. We further draw on multiple rounds of the HFPS to create variables on respondents’ attitudes towards the covid-19 emergency and how it has affected them and their families. These include willingness to be tested for covid-19, rating of government response, and whether households received any government assistance during the pandemic. The data as well as survey instruments and basic information documents are available publicly on the World Bank Microdata Library under the High-Frequency Phone Survey collection.[13]

### Sampling and sample representativeness

The samples for the HFPS were drawn from mobile phone numbers recorded during data collection for nationally representative face-to-face (FtF) household surveys implemented prior to the covid-19 pandemic with support from the World Bank Living Standards Measurement Study – Integrated Surveys on Agriculture (LSMS-ISA) program. Specifically, the Burkina Faso Enquête harmonisée sur les conditions de vie des ménages 2018/19, Ethiopia Socioeconomic Survey 2018/19, Malawi Integrated Household Panel Survey 2019, Mali Enquête harmonisée sur les conditions de vie des ménages 2018/19, Nigeria General Household Survey – Panel 2018/19, and Uganda National Panel Survey 2019/20. At least one phone number was recorded for all households with access to a phone, including through a contact person outside of the household, such as a friend or neighbor. For the HFPS, Ethiopia, Malawi, and Uganda attempted to contact all households with available phone numbers, while Burkina Faso, Nigeria and Mali selected a random sub-sample of phone numbers to call (Table A.1).

In the absence of universal access to a mobile phone, the HFPS households are likely to be selected samples of the nationally representative FtF survey samples of households.[14] The publicly available HFPS datasets therefore come with recalibrated household survey weights to counteract potential selection biases. The recalibration model takes advantage of the rich information on households with and without access to a phone from the pre-covid-19 FtF surveys [15,16] and follows a methodology proposed in a reference methodological paper on this subject.[17] The effectiveness of this weight recalibration in overcoming selection biases has been documented in HFPS data from four of the six countries we study, such that household-level estimates based on the HFPS data can be broadly considered representative at the national level.[14]

Further, the HFPS survey questions on vaccinations were asked only to the household’s main respondent, who had to be 15 or older, as it is impractical to interview all individuals in a household in a phone survey. The selection of main respondents was not randomized so that the group of respondents is likely not representative of the general population of adults at the individual level. Rather, respondents tend to be household heads or their spouses, better educated and slightly older, as a recent study documents.[18] An additional recalibration of survey weights for individual-level analysis improves representativeness, but cannot overcome respondent selection biases in all variables and increases the variance of the estimates.[18]

Considering these potential shortfalls in the representativeness of our data, we present our main results first with the standard HFPS household weights and then, as a robustness and sensitivity check for potential respondent selection biases, with the recalibrated individual-level weights. A summary of unweighted individual characteristics of FtF adults vis-à-vis HFPS adults is presented in Table A.3.

### Statistical analysis

The statistical analysis proceeds in several steps. First, we estimate the weighted mean of willingness to get vaccinated by country, and within countries by sex of respondent, residence (urban and rural) and income quintile, as well as reasons for vaccine hesitancy, using the recalibrated household weights. Second, we explore how individual and household characteristics, such as education and expenditure, correlate with the willingness to get vaccinated in a set of multivariate logit regressions, again using household weights. To assess how much the differences in the attributes of respondents vis-à-vis the general adult population may affect the representativeness of our results, we assess the sensitivity of the results to using individual-level recalibrated weights instead of the public-use household weights (see Discussion).[18]

### Patient and public involvement

This research did not involve consultation with patients or the public.

## Results

### Descriptive results

Overall, we find high levels of willingness to be vaccinated: Acceptance is estimated to be nearly universal in Ethiopia at 97.9% (95% confidence interval 97.2% to 98.6%) and very high in Nigeria (86.2%, 83.9% to 88.5%), Uganda (84.5%, 82.2% to 86.8%), Malawi (82.7%, 80.0% to 85.4%), and Burkina Faso (79.5%, 76.9% to 82.1%). In these countries at least four in five respondents would agree to be vaccinated if an approved vaccine was made available for free to them (Figure 1, Panel A). Acceptance is somewhat lower only in Mali where less than two thirds of respondents (64.5%, 61.3% to 67.8%) reported their willingness to be vaccinated. Notably, Mali is also the only country in which a non-negligible share (12.4%) is uncertain about their answer, a fact that accounts for the majority of the lower acceptance rates in Mali. For the following analysis, we focus on acceptance rates and do not distinguish between respondents rejecting to be vaccinated and those that are not sure. Pooling the data from all countries and weighting them by their respective popualtion sizes yields an overall mean acceptance rate of 87.6% (86.4% to 88.8%) across the six countries (Table A.2).

**Figure 1:**
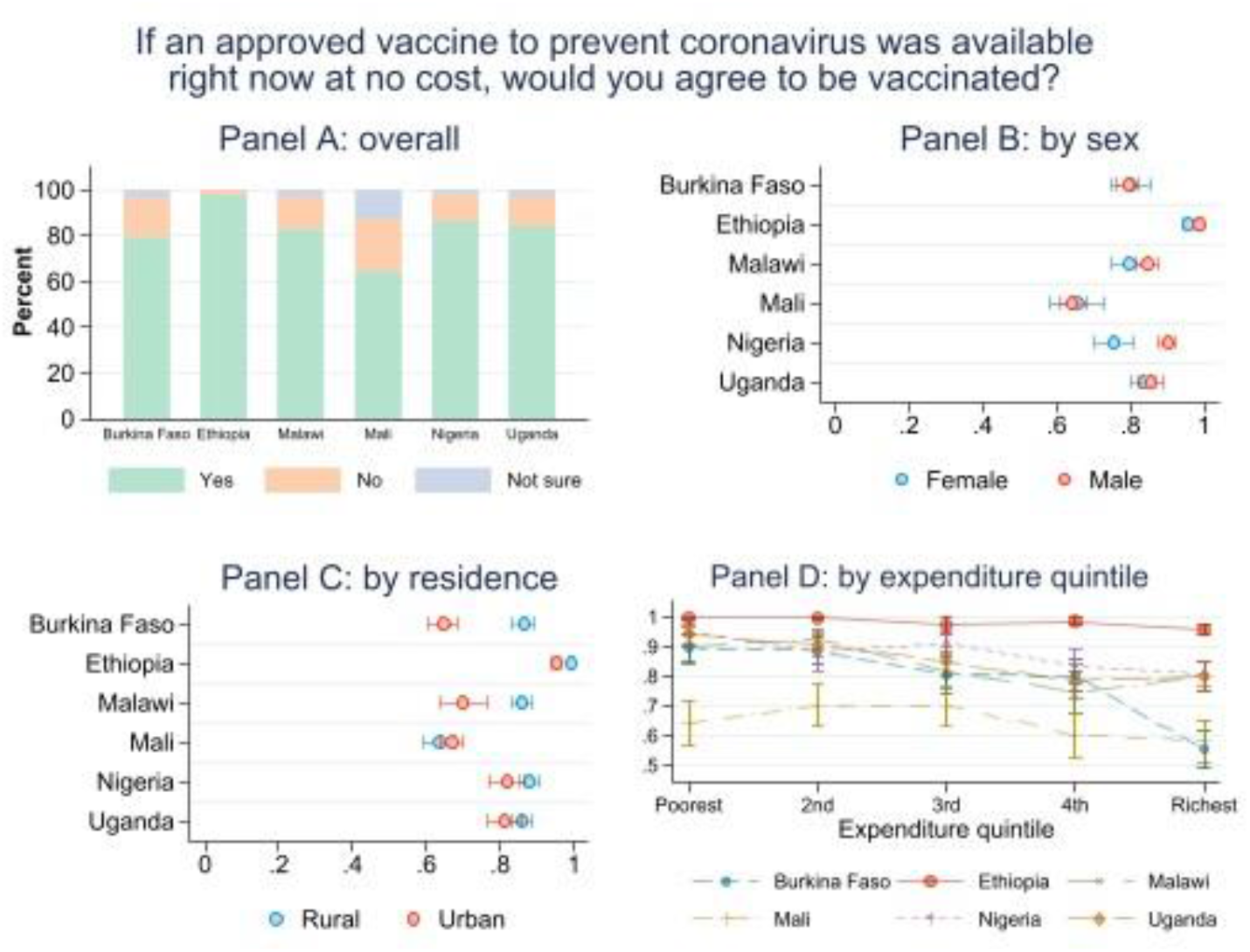
Vaccine hesitancy overall, by sex, by residence, and by expenditure

In Panel B of Figure 1, we disaggregate acceptance rates by respondent sex. Except for Nigeria where acceptance is statistically significantly higher among male than female respondents (90.1%, 87.6% to 92.6% and 75.7%, 70.4% to 80.9%, respectively) and in Ethiopia where there is a small, yet statistically significant difference (98.7%, 97.9% to 99.4% and 95.8%, 94.1% to 97.5%, respectively), we do not find answers to differ between men and women. Similarly, Panel C reports differences in willingness to be vaccinated against covid-19 between rural and urban areas with higher acceptance in rural areas in Burkina Faso, Ethiopia, and Malawi at the 95% confidence level. One distinct feature of our data is the ability to tap into the rich pre-covid-19 baseline data from the face-to-face household surveys that served as sampling frames. This way, we are able to determine respondent household’s position in the national expenditure distribution and disaggregated acceptance rates by expenditure quintile (Panel D). We generally find a downward sloping pattern in which acceptance is higher among poorer households and lowest among the richest households. This pattern is particularly evident in Burkina Faso but also noteworthy in Uganda and Nigeria. Conversely, acceptance is high throughout all expenditure quintiles in Ethiopia and not significantly lower for the richest two quintiles in Mali.

For those respondents who were hesitant to be vaccinated, the survey asked to provide a reason. From Figure 2, it is evident that safety concerns were paramount ranging from 53.7% (36.1% to 71.3%) of those reporting they would refuse to be vaccinated in Burkina Faso to 42.4% (33.8% to 50.9%) in Malawi, despite the wording of the questions making it explicit that the vaccine would be officially approved. Other notable reasons include worries about the potential side effects of the vaccine (between 55% (46.9% to 63.0%) in Uganda and 12.8% (9.1% to 16.5%) in Mali) and the belief not to be at risk of contracting the virus (34.2% (25.2% to 43.3%) in Nigeria to 0.1% (0% to 2.1%) in Burkina Faso).

**Figure 2:**
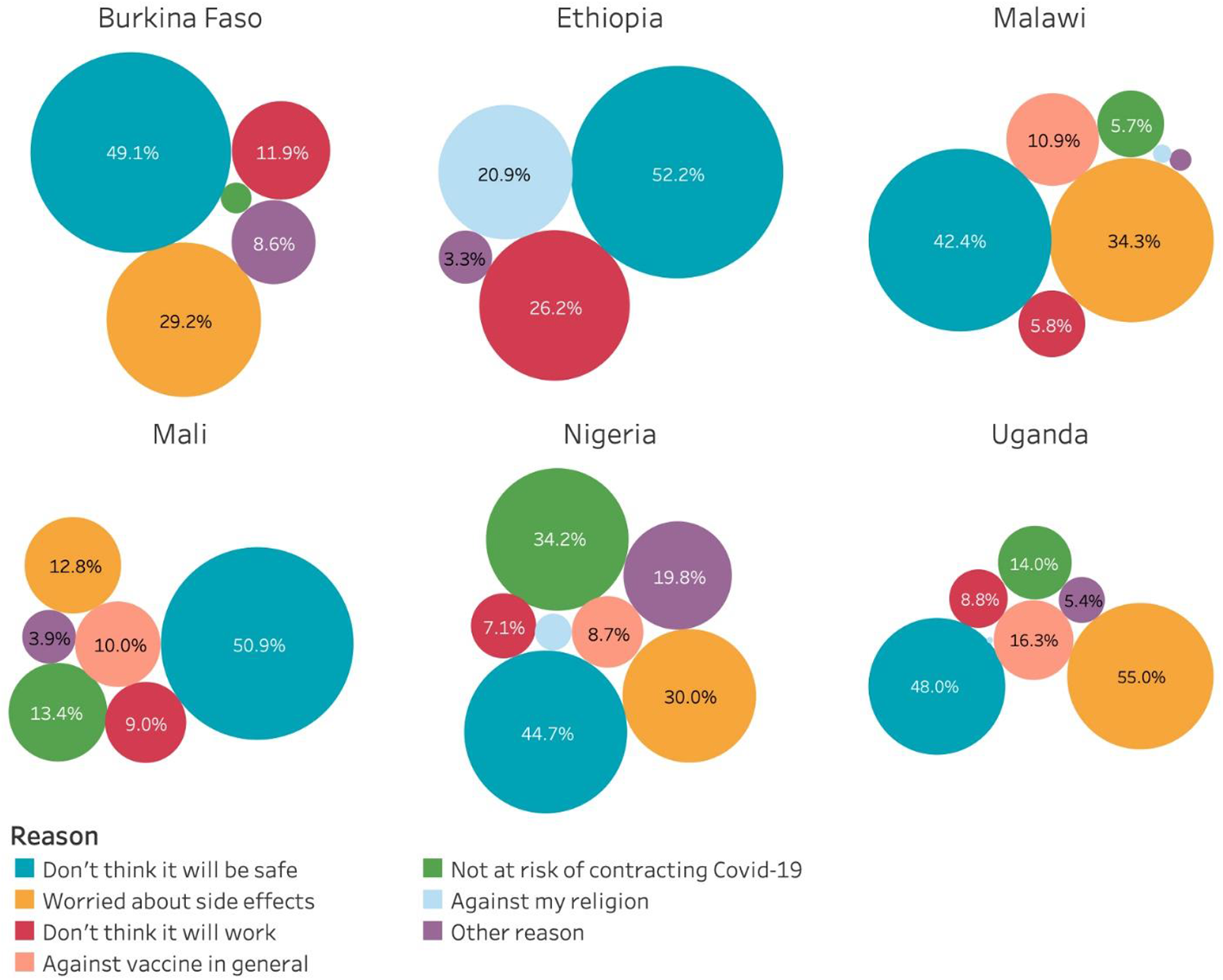
Reasons for covid-19 vaccine hesitancy

### Correlates of hesitancy

Exploiting the richness of our data, we can explore correlational patterns between willingness to be vaccinated for covid-19 and a large set of respondent and household characteristics. Table 1 confirms in a multivariate setting the patterns observed graphically earlier. In general, men tend to be more willing to take the vaccine, though we only detect a statistically significant effect in Nigeria. Respondents in urban areas are more skeptical with significant differences in Burkina Faso, Ethiopia, and Malawi.

**Table 1:**
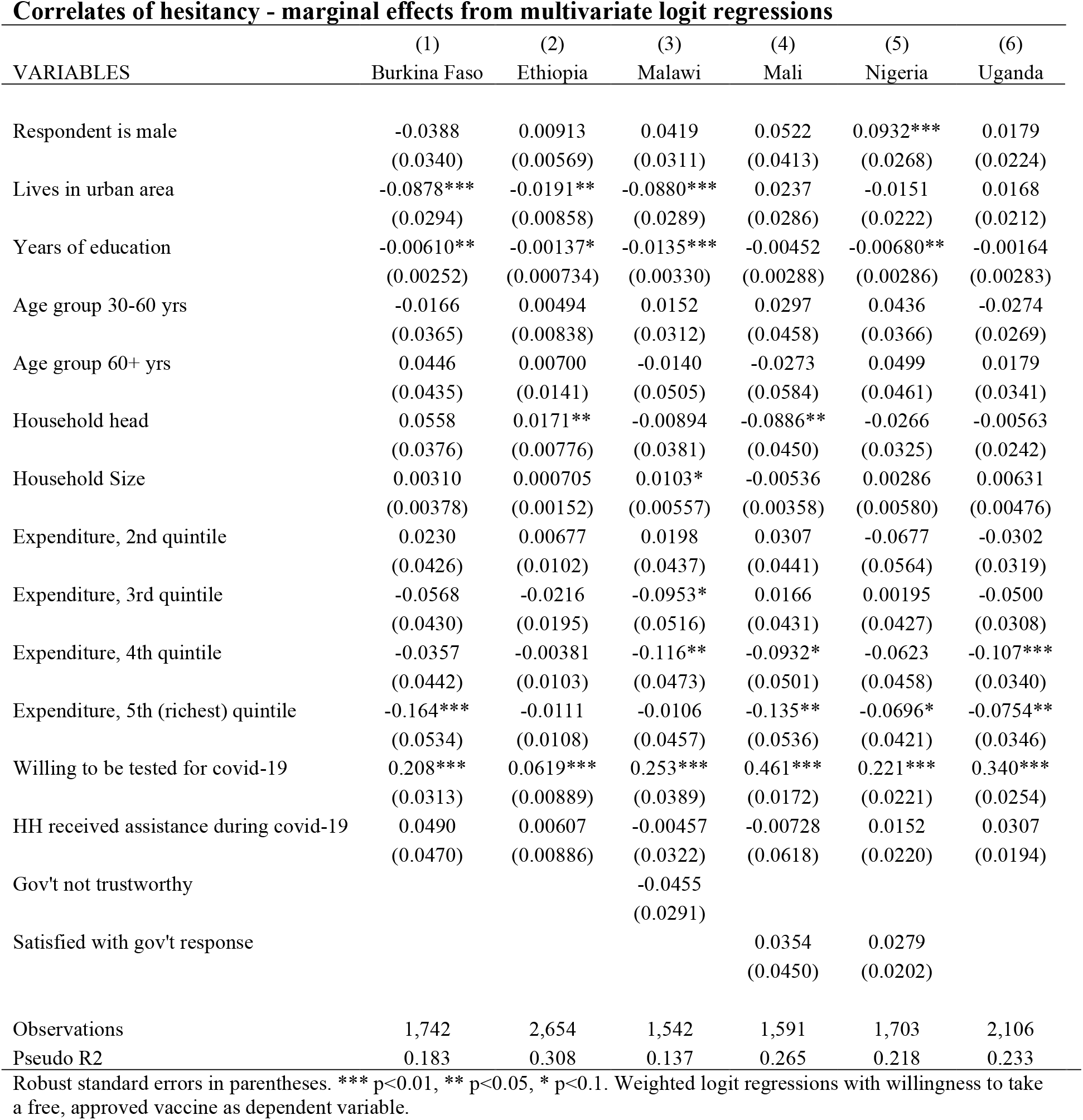
Correlates of hesitancy.

| **Correlates of hesitancy - marginal effects from multivariate logit regressions** |  |  |  |  |  |  |
| --- | --- | --- | --- | --- | --- | --- |
| VARIABLES | (1)<br>Burkina Faso | (2)<br>Ethiopia | (3)<br>Malawi | (4)<br>Mali | (5)<br>Nigeria | (6)<br>Uganda |
| Respondent is male | -0.0388<br>(0.0340) | 0.00913<br>(0.00569) | 0.0419<br>(0.0311) | 0.0522<br>(0.0413) | 0.0932***<br>(0.0268) | 0.0179<br>(0.0224) |
| Lives in urban area | -0.0878***<br>(0.0294) | -0.0191**<br>(0.00858) | -0.0880***<br>(0.0289) | 0.0237<br>(0.0286) | -0.0151<br>(0.0222) | 0.0168<br>(0.0212) |
| Years of education | -0.00610**<br>(0.00252) | -0.00137*<br>(0.000734) | -0.0135***<br>(0.00330) | -0.00452<br>(0.00288) | -0.00680**<br>(0.00286) | -0.00164<br>(0.00283) |
| Age group 30-60 yrs | -0.0166<br>(0.0365) | 0.00494<br>(0.00838) | 0.0152<br>(0.0312) | 0.0297<br>(0.0458) | 0.0436<br>(0.0366) | -0.0274<br>(0.0269) |
| Age group 60+ yrs | 0.0446<br>(0.0435) | 0.00700<br>(0.0141) | -0.0140<br>(0.0505) | -0.0273<br>(0.0584) | 0.0499<br>(0.0461) | 0.0179<br>(0.0341) |
| Household head | 0.0558<br>(0.0376) | 0.0171**<br>(0.00776) | -0.00894<br>(0.0381) | -0.0886**<br>(0.0450) | -0.0266<br>(0.0325) | -0.00563<br>(0.0242) |
| Household Size | 0.00310<br>(0.00378) | 0.000705<br>(0.00152) | 0.0103*<br>(0.00557) | -0.00536<br>(0.00358) | 0.00286<br>(0.00580) | 0.00631<br>(0.00476) |
| Expenditure, 2nd quintile | 0.0230<br>(0.0426) | 0.00677<br>(0.0102) | 0.0198<br>(0.0437) | 0.0307<br>(0.0441) | -0.0677<br>(0.0564) | -0.0302<br>(0.0319) |
| Expenditure, 3rd quintile | -0.0568<br>(0.0430) | -0.0216<br>(0.0195) | -0.0953*<br>(0.0516) | 0.0166<br>(0.0431) | 0.00195<br>(0.0427) | -0.0500<br>(0.0308) |
| Expenditure, 4th quintile | -0.0357<br>(0.0442) | -0.00381<br>(0.0103) | -0.116**<br>(0.0473) | -0.0932*<br>(0.0501) | -0.0623<br>(0.0458) | -0.107***<br>(0.0340) |
| Expenditure, 5th (richest) quintile | -0.164***<br>(0.0534) | -0.0111<br>(0.0108) | -0.0106<br>(0.0457) | -0.135**<br>(0.0536) | -0.0696*<br>(0.0421) | -0.0754**<br>(0.0346) |
| Willing to be tested for covid-19 | 0.208***<br>(0.0313) | 0.0619***<br>(0.00889) | 0.253***<br>(0.0389) | 0.461***<br>(0.0172) | 0.221***<br>(0.0221) | 0.340***<br>(0.0254) |
| HH received assistance during covid-19 | 0.0490<br>(0.0470) | 0.00607<br>(0.00886) | -0.00457<br>(0.0322) | -0.00728<br>(0.0618) | 0.0152<br>(0.0220) | 0.0307<br>(0.0194) |
| Gov't not trustworthy |  |  | -0.0455<br>(0.0291) |  |  |  |
| Satisfied with gov't response |  |  |  | 0.0354<br>(0.0450) | 0.0279<br>(0.0202) |  |
| Observations | 1,742 | 2,654 | 1,542 | 1,591 | 1,703 | 2,106 |
| Pseudo R2 | 0.183 | 0.308 | 0.137 | 0.265 | 0.218 | 0.233 |

Robust standard errors in parentheses. \*\*\* p<0.01, \*\* p<0.05, \* p<0.1. Weighted logit regressions with willingness to take a free, approved vaccine as dependent variable.

An association we observe across countries is between education and vaccine hesitancy. In Burkina Faso, Ethiopia, Malawi, and Nigeria, those with more years of education are significantly less willing to be vaccinated while coefficients signs point in the same direction in Mali and Uganda but are not significant. Lastly, we do not find a pattern according to respondents’ age and a mixed picture according to the role of the respondent in the household. Here, household heads which constitute the majority of our sample (see Table A.3) are more willing to be vaccinated in Ethiopia, more hesitant in Mali, and not significantly different from other household members in the remaining four cases.

A pattern we observe in several countries is a higher hesitancy toward the vaccine in richer households compared to the poorest expenditure quintile, with coefficients significant in Mali and Uganda for the richest two quintiles, for the top quintile in Burkina Faso and Nigeria, and the third and fourth quintiles in Malawi.

The data further show a strong association between vaccine hesitancy and the willingness to be tested for covid-19 suggesting similar underlying reasons for (or against) testing and getting vaccinated. Another hypothesis we explore is whether there is an association between the receipt of some assistance during the pandemic, e.g. through one of the large-scale social protection programs launched to combat the fallout of the pandemic, and willingness to receive a free, approved vaccine against covid-19. While coefficient signs mostly point to a positive association, the effect is not statistically significantly different from zero despite all countries in our sample launching some social protection response to the crisis.[19]

A last hypothesis we pursue concerns the relationship between trust in and satisfaction with the government and willingness to be vaccinated. For example, more skeptical individuals toward the government or those dissatisfied with its crisis management may be more reluctant to accept a state-provided vaccine.[20,21] We only have data on trust in the government’s crisis management in Malawi and satisfaction in Mali and Nigeria but find our hypotheses confirmed in bivariate logistic regressions (Table A.6). However, after controlling for all other factors in Table 1, coefficients are no longer significant.

## Discussion

### Principal findings

Our study uses cross-country comparable data from the national longitudinal high frequency phone surveys on covid-19 (HFPS) in six Sub-Saharan African countries to estimate acceptance rates for an approved, free covid-19 vaccine. By linking phone survey data to the nationally representative, large-scale, face-to-face household surveys that served as sampling frames for the HFPS, we provide robust estimates of the willingness to be vaccinated against covid-19 across a diverse set of demographics and respondent and household characteristics. Our headline results indicate high acceptance rates with at least four in five respondents signaling their willingness to be vaccinated in all but one of the countries studied. There is cross-country variation, with willingness to be vaccinated ranging from 64.5% in Mali (61.3% to 67.8%), where a further 12.4% are yet undecided, to near universal acceptance in Ethiopia (97.9%, 97.2% to 98.6%). We also find little evidence for systematic differences in vaccine hesitancy across gender or age but some notable clusters of hesitancy in urban areas, among those better educated, and in richer households. Across countries, safety concerns about the vaccine in general and its side effects specifically emerged as the primary reservations toward a covid-19 vaccine.

### Strengths and comparison with other studies

In relation to the existing literature, our study has several advantages in the domains of coverage and sample selection, richness of the data, and analytical depth. In terms of coverage, our study focuses on a region with yet scant evidence on willingness to be vaccinated against covid-19. Importantly, the data we assemble are cross-country comparable and without exception based on large, nationally representative sampling frames from pre-covid, face-to-face household surveys. This allows us to calibrate survey weights that adjust for the coverage and non-response biases that plague other studies at a time where regular face-to-face data collection nearly came to a complete halt.[22–24] Furthermore, our study is unique in that the pre-existing survey data from which our samples are drawn allow us to tap into a rich set of baseline individual and household characteristics. This facilitates a more rigorous analysis and disaggregation of acceptance rates, including, for instance, households’ position in the national (pre-crisis) expenditure distribution, than has been possible previously. Our study thus stands out as a fully cross-country comparable, multivariate, and inferential analysis of vaccine hesitancy on the African continent.

Compared to our study, previous studies that analyze covid-19 vaccine acceptance rates and the reasons for refusal predominantly (i) focus on middle- and high-income countries, (ii) represent single-country or not cross-country comparable samples, (iii) study particular subpopulations such as university students, healthcare workers, or participants of unrelated pre-covid studies, (iv) rely on non-random sampling, or (v) can only draw on a small set of demographics and characteristics to disaggregate their analysis.[18,25–35]

A cross-country study of 19 countries with samples obtained from commercial online panel providers found 71.5% of respondents willing to be vaccinated against covid-19 with a rate of 65.2% in Nigeria as the only low-income country.[36] Similarly, a literature review of 31 studies covering 33 countries found covid-19 vaccine acceptance rates at or above 70% among studies focusing on the general population but also large regional and intra-regional differences and a dearth of evidence particularly from Sub-Saharan Africa.[37]

Few studies explicitly focus on acceptance rates in the poorest countries. Assembling an amalgamation of data samples with different sources, sampling methodologies, and coverage, one study finds generally high acceptance rates in ten LMICs in Asia, Africa, and South America.[38] Among countries also covered in our study, acceptance rates are lower than what we find in Burkina Faso (66.5%, national phone sample obtained by random digit dialing) and Nigeria (76.2%, random sample of residents of one state from telephone list), close in urban Uganda (76.5%, random sample of households in Kampala), and very similar in rural Uganda (85.8%, non-random sample of women in 13 districts).[38] However, in those studies the ability to correct for various sample selection biases is likely limited in the absence of baseline nationally representative sampling frames. Another study in 15 African countries, including Burkina Faso, Malawi, Nigeria, Uganda (all face-to-face interviewing), and Ethiopia (telephone survey) found estimates for the countries also covered in our study mostly in the vicinity of our weighted figures (Ethiopia 94%, Uganda 87.5%, Burkina Faso 86%, Malawi 82.7%, Nigeria 76%).[39,40] Notably, average sample sizes are roughly half of our study’s and there is a lack of analytical detail impeding a robust inference of cross-country and -demographic findings. Lastly, our results contrast with a recent Afrobarometer study based on in-person interviews in five West African countries, none of which are also included in our sample. In relation to our study, the study’s analysis is purely descriptive and reports low acceptance rates of 40% on average.[21] Similar to what we find, the study cites trust in vaccine safety as the key driver of hesitancy, a result that is further corroborated in the literature.[21,28,36,38,40] Furthermore, the small systematic differences we observe according to gender and age are in line with most previous findings in low-income countries as is a tendency for higher acceptance in rural areas.[21,36,38,40] As with overall acceptance rates, our finding of higher hesitancy among the more educated is in line with two of the studies covering Sub-Saharan Africa [38,39] while another study finds mixed evidence.[21] No other study we are aware of across LMICs (and in fact few across countries of any income classification) assess vaccine acceptance according to economic status in a manner comparable with the expenditure data we can access from the pre-covid-19 sampling frames.

## Limitations of this study

This study uses data from high-frequency phone surveys (HFPS) with national coverage along with sampling weights specifically recalibrated for the phone surveys to be nationally representative. The weights were shown to be effective at achieving nationally representative estimates at the household level.[14] However, willingness to be vaccinated is primarily not a household-level attribute but an individual decision, though it is reasonable to expect considerable intra-household correlation in attitudes towards vaccination. Phone survey respondents in our data are not specifically selected to be representative of all individuals and that may limit the population-level representativeness of the results we report. Recalibration of survey weights at the individual level can partially but not fully address this concern.[18] To gauge how sensitive our results are to respondent selection biases in individual-level data, we compare the estimates with household-level weights to the same estimates with individual-level weights. In this test, large deviations between those two sets of estimates would indicate that selection biases affect our results in a fundamental way. However, we find only limited change in the estimates, regardless of which weight is used: in over three quarters of cases, deviations do not exceed two percentage points, including the headline findings on willingness to be vaccinated in all study countries, and we find only three instances where the differences exceed five points (Variable ‘Q5’ 5.8% in Burkina Faso; ‘Q4’ 6.2% in Mali; ‘Q3’ 6.0% in Nigeria; Table A.4). The results from the multivariate logit regression model are also robust to the use of household or individual-level weights. This is true especially of the cross-country findings for urban and richer households and willingness to get tested for covid-19: the point estimates tend to be slightly larger when using individual-level compared to household-level weights but they are of comparable magnitude and in most cases retain statistical significance (Table A.5). In contrast, the findings on education have similar point estimates whether we use household or individual-level weights, but with individual-level weights they retain statistical significance only in Malawi. This is likely because individual-level weight recalibration increases the variance of the estimates. All in all, we take this set of tests to indicate the robustness of our results to concerns around respondent selection.

Another potential limitation is the possible malleability of attitudes towards vaccinations, which may have changed, and continue to change, in light of the development of various vaccines and the relative success these appear to have in stemming the pandemic in other parts of the world. Future research is needed to determine evolving attitudes in Africa towards being vaccinated and their interactions with vaccine supply and availability.

## Conclusions and policy implications

Our headline results of high vaccine acceptance in a cross-country comparable sample of six Sub-Saharan African countries suggests that limited supply, not inadequate demand, is likely to present the key bottleneck to reaching high covid-19 vaccine coverage in the region. As willingness to be vaccinated does not automatically translate into vaccine-seeking behavior, public authorities need to turn intent into effective demand as vaccine rollout progresses.[41] For this, our study identifies some indicative pockets of hesitancy, particularly in Mali, urban Burkina Faso and Malawi, among women in Nigeria, and for richer households and those with more education. As many of these population groups are easier to reach early in the vaccine rollout process, yet are also better reachable through targeted communication, campaigns that raise acceptance for a covid-19 vaccine in these clusters will be key. These should focus on resolving concerns about side effects and bolster confidence in the safety of approved covid-19 vaccines in order to reach mass coverage and end the pandemic swiftly and everywhere.

## Supporting information

STROBE Checklist

## Data Availability

The data are available from the World Bank's Microdata Library, High Frequency Phone Survey Catalog https://microdata.worldbank.org/index.php/catalog/hfps) as well as LSMS catalog (https://microdata.worldbank.org/index.php/catalog/lsms).

https://microdata.worldbank.org/index.php/catalog/lsms

## Contributors

All authors had full access to all of the data in the study and take responsibility for the integrity of the data and the accuracy of the data analysis. All authors were responsible for its conception and design. YM, PW, and SK were responsible for the preparation and analysis of the data and all authors contributed to their interpretation. YM and PW drafted the manuscript and AZ made critical revision of the manuscript for important intellectual content. SK and AZ conceptualized an early version. AZ is the guarantor. The corresponding author attests that all listed authors meet authorship criteria and that no others meeting criteria have been omitted.

## Ethics approval

Each phone survey was implemented by the respective national statistical office (NSO), except for Ethiopia where a private firm was the implementing agency. In Burkina Faso, Malawi, Mali, Nigeria, and Uganda, the NSO conducts the survey as the sole official statistical authority in the country and in accordance with the respective National Statistical Act, which exempts the NSO from institutional ethics approvals. Informed consent was received from all survey respondents in each country. The World Bank does not require institutional ethics approval for household surveys that are partly or fully financed by the World Bank, including the national phone surveys in Burkina Faso, Ethiopia, Malawi, Mali, Nigeria, and Uganda that inform our research.

## Data Sharing

The data are available from the World Bank’s Microdata Library, High Frequency Phone Survey Catalog (https://microdata.worldbank.org/index.php/catalog/hfps) as well as LSMS catalog (https://microdata.worldbank.org/index.php/catalog/lsms).

## Transparency Statement

The authors affirm that the manuscript is an honest, accurate, and transparent account of the study being reported; that no important aspects of the study have been omitted; and that any discrepancies from the study as planned have been explained.

## Dissemination to participants and related patient and public communities

Our research uses data from a large, anonymized sample of participants and there are thus no plans to disseminate the research to specific participants or patients beyond publishing it.

## Licensing

The Corresponding Author is aware that the World Bank external publishing department has recently agreed to grant to BMJ Publishing Group Limited a worldwide, non-exclusive license to publish an article produced by World Bank staff and consultants, and is committed to facilitate the conclusion of a similar agreement should this manuscript be accepted for publication.

## Appendix

**Table A.1.** Household selection, number of households

| *Panel A. Households (N)* | **Burkina**<br>**Faso** | **Ethiopia** | **Malawi** | **Mali** | **Nigeria** | **Uganda** |
| --- | --- | --- | --- | --- | --- | --- |
| Pre-COVID-19 FtF survey | 7,010 | 6,770 | 3,181 | 7,000 | 4,976 | 3,098 |
| With phone numbers | 6,877 | 5,374 | 2,337 | 6,300 | 4,934 | 2,386 |
| Attempted to contact | 2,500 | 5,374 | 2,337 | 2,270 | 3,000 | 2,386 |
| Reached | 2,062 | 3,357 | 1,743 | 2,270 | 2,057 | 2,246 |
| Phone interviews completed | 1,945 | 2,701 | 1,589 | 1,765 | 1,766 | 2,129 |

**Table A.2.** Willingness to get vaccinated by country and pooled, mean and 95% confidence interval

|  | Mean | Lower Bound | Upper Bound | N |
| --- | --- | --- | --- | --- |
| Burkina Faso, weighted | 79.6% | 77.0% | 82.3% | 1,945 |
| Ethiopia, weighted | 97.9% | 97.2% | 98.6% | 2,701 |
| Malawi, weighted | 82.7% | 80.0% | 85.4% | 1,589 |
| Mali, weighted | 64.5% | 61.3% | 67.8% | 1,765 |
| Nigeria, weighted | 86.2% | 83.9% | 88.5% | 1,766 |
| Uganda, weighted | 84.5% | 82.2% | 86.8% | 2,129 |
| Pooled, weighted | 87.6% | 86.4% | 88.8% | 11,895 |
| Pooled, unweighted | 82.4% | 81.7% | 83.1% |  |

**Table A. 3.** Unweighted descriptive statistics in the baseline F2F surveys and the HFPS.

|  | Burkina Faso |  | Ethiopia |  | Malawi |  | Mali |  | Nigeria |  | Uganda |  |
| --- | --- | --- | --- | --- | --- | --- | --- | --- | --- | --- | --- | --- |
|  | F2F | HFPS | F2F | HFPS | F2F | HFPS | F2F | HFPS | F2F | HFPS | F2F | HFPS |
| Men | 45.5 | 78.7 | 47.3 | 62.7 | 47.6 | 59.8 | 47.4 | 78.9 | 48.3 | 72.3 | 48.2 | 50.7 |
| HH heads | 28.7 | 86.1 | 38.5 | 82.5 | 37.0 | 74.5 | 27.1 | 82.3 | 32.7 | 79.3 | 35.1 | 74.5 |
| Urban | 44.9 | 72.0 | 54.0 | 71.9 | 26.0 | 36.8 | 41.7 | 66.3 | 32.0 | 39.3 | 24.2 | 25.8 |
| Primary education | 26.1 | 32.4 | 34.6 | 52.9 | 37.2 | 50.2 | 20.2 | 30.9 | 66.5 | 75.1 | 47.8 | 51.1 |
| < 30 years | 44.1 | 11.3 | 49.6 | 29.4 | 52.9 | 27.8 | 42.6 | 13.8 | 42.5 | 13.5 | 48.2 | 15.3 |
| 30 - 60 years | 45.7 | 72.2 | 42.1 | 62.4 | 38.2 | 62.7 | 45.7 | 70.4 | 44.9 | 69.0 | 40.0 | 66.2 |
| 60+ years | 10.1 | 16.5 | 8.3 | 8.3 | 8.9 | 9.5 | 11.7 | 15.9 | 12.6 | 17.5 | 11.8 | 18.6 |
| Quintile 1 (poorest) | 18.7 | 14.0 | 11.4 | 4.6 | 15.7 | 8.7 | 18.2 | 14.6 | 12.7 | 9.5 | 18.2 | 15.8 |
| Quintile 2 | 18.9 | 15.6 | 13.8 | 9.5 | 17.6 | 14.6 | 18.9 | 16.4 | 14.4 | 13.1 | 19.0 | 18.5 |
| Quintile 3 | 19.9 | 17.9 | 15.0 | 12.4 | 17.4 | 16.1 | 21.0 | 19.0 | 19.2 | 18.7 | 20.4 | 20.7 |
| Quintile 4 | 21.3 | 22.5 | 20.3 | 22.0 | 22.2 | 23.5 | 20.7 | 22.2 | 21.4 | 21.9 | 21.7 | 22.5 |
| Quintile 5 (richest) | 21.2 | 30.0 | 39.4 | 51.5 | 27.1 | 37.1 | 21.2 | 27.8 | 32.3 | 36.9 | 20.7 | 22.5 |
| **Observations** | 24,396 | 1,742 | 17,563 | 2,701 | 8,588 | 1,589 | 24,394 | 1,765 | 15,230 | 1,766 | 8,763 | 2,129 |

Note: The table shows descriptive statistics and sample sizes for the face-to-face (F2F) pre-Covid samples of the general adult population (>15 years) and the high-frequency phone surveys (HFPS) during the pandemic. All values except observation numbers in percent.

**Table A.4.**
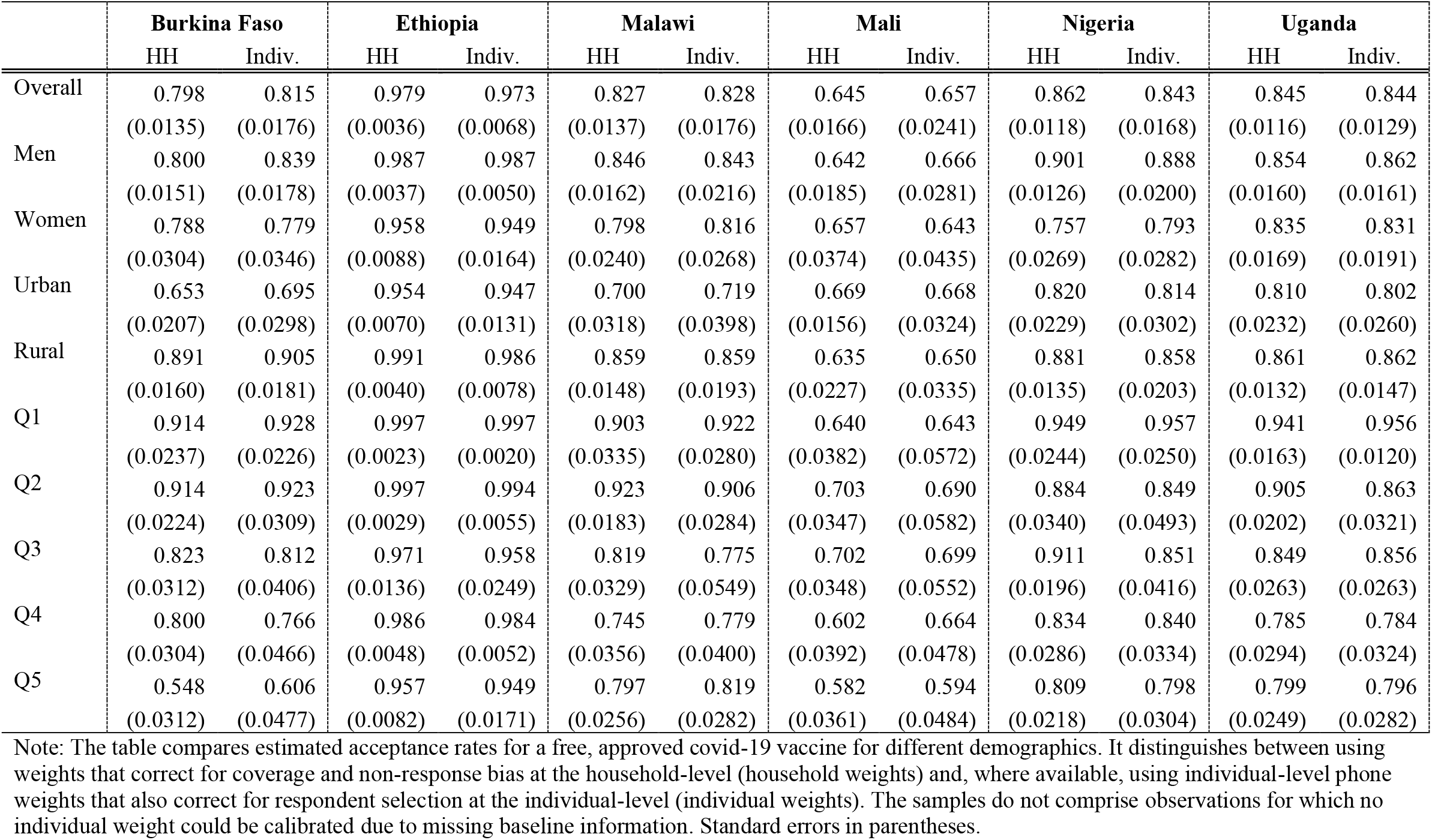
Estimated acceptance rates using household or individual weights.

|  | Burkina Faso |  | Ethiopia |  | Malawi |  | Mali |  | Nigeria |  | Uganda |  |
| --- | --- | --- | --- | --- | --- | --- | --- | --- | --- | --- | --- | --- |
|  | HH | Indiv. | HH | Indiv. | HH | Indiv. | HH | Indiv. | HH | Indiv. | HH | Indiv. |
| Overall | 0.798<br>(0.0135) | 0.815<br>(0.0176) | 0.979<br>(0.0036) | 0.973<br>(0.0068) | 0.827<br>(0.0137) | 0.828<br>(0.0176) | 0.645<br>(0.0166) | 0.657<br>(0.0241) | 0.862<br>(0.0118) | 0.843<br>(0.0168) | 0.845<br>(0.0116) | 0.844<br>(0.0129) |
| Men | 0.800<br>(0.0151) | 0.839<br>(0.0178) | 0.987<br>(0.0037) | 0.987<br>(0.0050) | 0.846<br>(0.0162) | 0.843<br>(0.0216) | 0.642<br>(0.0185) | 0.666<br>(0.0281) | 0.901<br>(0.0126) | 0.888<br>(0.0200) | 0.854<br>(0.0160) | 0.862<br>(0.0161) |
| Women | 0.788<br>(0.0304) | 0.779<br>(0.0346) | 0.958<br>(0.0088) | 0.949<br>(0.0164) | 0.798<br>(0.0240) | 0.816<br>(0.0268) | 0.657<br>(0.0374) | 0.643<br>(0.0435) | 0.757<br>(0.0269) | 0.793<br>(0.0282) | 0.835<br>(0.0169) | 0.831<br>(0.0191) |
| Urban | 0.653<br>(0.0207) | 0.695<br>(0.0298) | 0.954<br>(0.0070) | 0.947<br>(0.0131) | 0.700<br>(0.0318) | 0.719<br>(0.0398) | 0.669<br>(0.0156) | 0.668<br>(0.0324) | 0.820<br>(0.0229) | 0.814<br>(0.0302) | 0.810<br>(0.0232) | 0.802<br>(0.0260) |
| Rural | 0.891<br>(0.0160) | 0.905<br>(0.0181) | 0.991<br>(0.0040) | 0.986<br>(0.0078) | 0.859<br>(0.0148) | 0.859<br>(0.0193) | 0.635<br>(0.0227) | 0.650<br>(0.0335) | 0.881<br>(0.0135) | 0.858<br>(0.0203) | 0.861<br>(0.0132) | 0.862<br>(0.0147) |
| Q1 | 0.914<br>(0.0237) | 0.928<br>(0.0226) | 0.997<br>(0.0023) | 0.997<br>(0.0020) | 0.903<br>(0.0335) | 0.922<br>(0.0280) | 0.640<br>(0.0382) | 0.643<br>(0.0572) | 0.949<br>(0.0244) | 0.957<br>(0.0250) | 0.941<br>(0.0163) | 0.956<br>(0.0120) |
| Q2 | 0.914<br>(0.0224) | 0.923<br>(0.0309) | 0.997<br>(0.0029) | 0.994<br>(0.0055) | 0.923<br>(0.0183) | 0.906<br>(0.0284) | 0.703<br>(0.0347) | 0.690<br>(0.0582) | 0.884<br>(0.0340) | 0.849<br>(0.0493) | 0.905<br>(0.0202) | 0.863<br>(0.0321) |
| Q3 | 0.823<br>(0.0312) | 0.812<br>(0.0406) | 0.971<br>(0.0136) | 0.958<br>(0.0249) | 0.819<br>(0.0329) | 0.775<br>(0.0549) | 0.702<br>(0.0348) | 0.699<br>(0.0552) | 0.911<br>(0.0196) | 0.851<br>(0.0416) | 0.849<br>(0.0263) | 0.856<br>(0.0263) |
| Q4 | 0.800<br>(0.0304) | 0.766<br>(0.0466) | 0.986<br>(0.0048) | 0.984<br>(0.0052) | 0.745<br>(0.0356) | 0.779<br>(0.0400) | 0.602<br>(0.0392) | 0.664<br>(0.0478) | 0.834<br>(0.0286) | 0.840<br>(0.0334) | 0.785<br>(0.0294) | 0.784<br>(0.0324) |
| Q5 | 0.548<br>(0.0312) | 0.606<br>(0.0477) | 0.957<br>(0.0082) | 0.949<br>(0.0171) | 0.797<br>(0.0256) | 0.819<br>(0.0282) | 0.582<br>(0.0361) | 0.594<br>(0.0484) | 0.809<br>(0.0218) | 0.798<br>(0.0304) | 0.799<br>(0.0249) | 0.796<br>(0.0282) |

Note: The table compares estimated acceptance rates for a free, approved covid-19 vaccine for different demographics. It distinguishes between using weights that correct for coverage and non-response bias at the household-level (household weights) and, where available, using individual-level phone weights that also correct for respondent selection at the individual-level (individual weights). The samples do not comprise observations for which no individual weight could be calibrated due to missing baseline information. Standard errors in parentheses.

**Table A.5.** Logistic regression on the correlates of hesitancy using individual-level survey weights, marginal effects.

| VARIABLES | (1)<br>Burkina<br>Faso | (2)<br>Ethiopia | (3)<br>Malawi | (4)<br>Mali | (5)<br>Nigeria | (6)<br>Uganda |
| --- | --- | --- | --- | --- | --- | --- |
| Respondent is male | 0.0265<br>(0.0351) | 0.0125<br>(0.00942) | 0.0272<br>(0.0371) | 0.116**<br>(0.0494) | 0.0307<br>(0.0406) | 0.00464<br>(0.0244) |
| Lives in urban area | -0.0750**<br>(0.0323) | -0.0236**<br>(0.0116) | -0.0930***<br>(0.0348) | -0.0221<br>(0.0388) | -0.00481<br>(0.0370) | 0.00853<br>(0.0223) |
| Years of education | -0.00462<br>(0.00293) | -0.00154<br>(0.000938) | -0.0123***<br>(0.00417) | -0.00337<br>(0.00356) | -0.00657<br>(0.00410) | -0.00261<br>(0.00247) |
| Age group, 30-60 yrs | 0.0354<br>(0.0490) | -0.00114<br>(0.00984) | -0.00707<br>(0.0387) | -0.0117<br>(0.0476) | -0.0124<br>(0.0356) | -0.0340<br>(0.0294) |
| Age group, 60+ yrs | 0.124**<br>(0.0491) | 0.00134<br>(0.0196) | -0.0331<br>(0.0720) | -0.0471<br>(0.0612) | -0.0321<br>(0.0628) | 0.0134<br>(0.0317) |
| Household head | -0.00843<br>(0.0401) | 0.0226*<br>(0.0119) | -0.00400<br>(0.0385) | -0.0972*<br>(0.0549) | 0.0333<br>(0.0415) | 0.0204<br>(0.0267) |
| Household Size | 0.00287<br>(0.00510) | 0.00186<br>(0.00181) | 0.00971<br>(0.00842) | -0.00534*<br>(0.00322) | 0.00834<br>(0.00591) | 0.00268<br>(0.00564) |
| Expenditure, 2nd quintile | 0.00107<br>(0.0502) | 0.00591<br>(0.0131) | -0.0203<br>(0.0469) | 0.0270<br>(0.0638) | -0.0994<br>(0.0757) | -0.0871***<br>(0.0323) |
| Expenditure, 3rd quintile | -0.0660<br>(0.0428) | -0.0277<br>(0.0193) | -0.178**<br>(0.0712) | 0.0325<br>(0.0537) | -0.0371<br>(0.0666) | -0.0813***<br>(0.0287) |
| Expenditure, 4th quintile | -0.0921**<br>(0.0466) | -0.00480<br>(0.0118) | -0.118**<br>(0.0522) | -0.0504<br>(0.0595) | -0.0428<br>(0.0588) | -0.122***<br>(0.0316) |
| Expenditure, 5th (richest) quintile | -0.123**<br>(0.0528) | -0.0157<br>(0.0126) | -0.0143<br>(0.0461) | -0.115*<br>(0.0684) | -0.0679<br>(0.0631) | -0.103***<br>(0.0317) |
| Willing to be tested for covid-19 | 0.271***<br>(0.0287) | 0.0839***<br>(0.0149) | 0.249***<br>(0.0440) | 0.468***<br>(0.0171) | 0.224***<br>(0.0302) | 0.348***<br>(0.0287) |
| HH received assistance during covid-19 | 0.0840*<br>(0.0505) | 0.0177<br>(0.0127) | -0.0412<br>(0.0455) | 0.0406<br>(0.0598) | -0.00112<br>(0.0328) | 0.0597***<br>(0.0214) |
| Gov't not trustworthy |  |  | -0.0408<br>(0.0322) |  |  |  |
| Satisfied with gov't response |  |  |  | 0.0356<br>(0.0446) | 0.0235<br>(0.0325) |  |
| Observations | 1,738 | 2,654 | 1,542 | 1,590 | 1,700 | 2,106 |
| Pseudo R2 | 0.225 | 0.357 | 0.136 | 0.325 | 0.162 | 0.238 |

Robust standard errors in parentheses. \*\*\* p<0.01, \*\* p<0.05, \* p<0.1. Weighted logit regressions with willingness to take a free, approved covid-19 vaccine as dependent variable.

**Table A.6:** Bivariate logistic regression on government trust and satisfaction using household-level survey weights.

| VARIABLES | (1)<br>Malawi | (2)<br>Mali | (3)<br>Nigeria |
| --- | --- | --- | --- |
| Government not trustworthy | -0.0638**<br>(0.0322) |  |  |
| Satisfied with government response |  | 0.161***<br>(0.0552) | 0.0568**<br>(0.0232) |
| Observations | 1,589 | 1,631 | 1,766 |
| Controls | YES | YES | YES |
| Pseudo R2 | 0.00574 | 0.00742 | 0.00847 |
| Robust standard errors in parentheses. *** p<0.01, ** p<0.05, * p<0.1 |  |  |  |

